# Profile of speech and cognitive impairments in a patient with moyamoya disease: a longitudinal case study

**DOI:** 10.1101/2023.08.29.23294546

**Authors:** A. Pashkov, E. Filimonova, A. Martirosyan, K. Ovsyannikov, G. Gunenko, G. Moysak, J. Rzaev

## Abstract

Moyamoya is a rare chronic brain vascular disease with a set of potential life-threatening consequences due to a high probability of stroke occurrence. Patients suffering from this disease are often presented with diverse and severe sensory, motor and cognitive deficits, which often lead to general inability to work and, in the most severe cases, to complete loss of self-serving skills and gross disturbance of vital homeostatic functions. Here we present a case of a patient with moyamoya disease, who has been dynamically observed over more than 4 years after the first manifestation of her symptoms. We have been tracking the appearance of patient’s new symptoms and resolution of existing ones after neurosurgical treatment (revascularization surgery) with in-depth neurological examination, neuropsychological assessment and fMRI/DTI sessions. This study represents one of the first attempts to rigorously estimate the patterns of deficits at different levels, connecting neural circuits affected by the disease to behavior. Our findings lend further support to the idea that revascularization surgery can improve cognitive performance and highlights the importance of long-term follow-up in understanding the effects of surgery on brain function.

## 1. Introduction

Moyamoya disease is a rare and chronic brain vascular disorder characterized by a gradual narrowing of the intracranial segments of the internal carotid artery (ICA) and the initial sections of the anterior and middle cerebral arteries, which can eventually lead to their occlusion. An identifying feature of the disease is the development of a network of intermingled vessels on the base of the brain, which appears as a light haze in angiogram images (Kuroda and Houkin, 2008). The progressive deterioration of blood supply conditions can result in a range of cognitive, emotional, and behavioral impairments, often accompanied by severe neurological deficits in these patients (Chan et al, 2023; Richards et al, 2019). Verbal memory, processing speed and executive functions are reportedly the most affected mental facilities, causing a significant drop in patient’s life quality, ability to work and live independently (Festa et al, 2010; Araki et al, 2014; Shi et al, 2020). In a study by Karzmark and colleagues, approximately one-third of the tested patients exhibited cognitive deficits, with disorders of executive functioning being the most severely affected cognitive domain, and perception being the least affected (Karzmark et al, 2008). These findings are consistent with those of a study conducted by Sun and colleagues, which showed that 31.5% (29 out of 92) of patients experienced cognitive dysfunctions (Sun et al, 2022). These behavioral dysfunctions are intricately linked to specific neural circuits or brain networks that support complex emotional and cognitive abilities, as well as a sophisticated interplay between them (Wang et al, 2023; Lei et al, 2020).

At the same time, speech impairments are rarely reported in patients with MMD, and few studies dedicated to examining speech in these patients are found in the international databases of scientific publications. Importantly, as the disease progresses and blood supply to the brain becomes increasingly compromised, patients may experience a range of speech-related issues, including difficulties with articulation, fluency, and language comprehension (Esin et al, 2016; McPherson and Leung, 2006). Recent studies have shown that progression of the MMD can cause damage to the brain regions responsible for speech and language, leading to a variety of dysfunctions. In particular, the disease has been associated with damage to the frontal and temporal lobes, which play key roles in speech production and comprehension (Hu et al, 2022; Saito et al, 2021). The study by Zuo and colleagues showed that nearly 30% of patients (in a sample of 142 individuals) with moyamoya disease had speech impairments in the form of dysarthria and aphasia (Zuo et al, 2023).

Currently, magnetic resonance imaging (MRI) and its modalities, such as MR angiography, MR perfusion, MR morphometry, functional MRI, diffusion tensor imaging, and magnetic resonance spectroscopy, play a leading role in diagnosing MMD progression, its association with neurological and behavioral deficits, and the degree of perfusion recovery after surgical treatment. This array of diverse neuroimaging techniques is now commonly utilized in clinical practice to aid clinicians in differential diagnostics and in determining an appropriate treatment plan for their patients (Lehman et al, 2019; Zhang et al, 2022).

Functional magnetic resonance imaging (fMRI) has also been used to investigate the neural basis of cognitive impairment in patients with moyamoya disease. Several studies have demonstrated altered brain activity in patients with moyamoya disease compared to healthy controls using fMRI (Kazumata et al., 2017; Lei et al., 2020). One of the main findings of fMRI studies in moyamoya disease is altered functional connectivity in the three canonical resting state networks, namely, default mode network (DMN), a network of brain regions that are active when the brain is at rest and not engaged in a specific task; salience network (SN), engaged in detecting salient or important external and internal events; and executive control network, regulating the performance of cognitive demanding tasks. These patterns of altered connectivity have been associated with cognitive impairment in patients with moyamoya disease (Lei et al., 2014).

However, due to the vascular origin of the disease and the heavy reliance on BOLD signals in fMRI paradigms to indirectly infer underlying neural tissue activity, caution should be exercised in readily trusting the obtained results. These results may be partially explained by the perfusion delay factor (Kazumata et al., 2017). Nevertheless, the findings are promising and support the feasibility of using fMRI to predict post-surgical cognitive improvement in patients (Lei et al., 2017; Gao et al, 2022).

Diffusion tensor imaging (DTI) is another imaging technique that has been extensively utilized to investigate structural changes in the brains of patients with moyamoya disease (Hao et al, 2022). DTI measures the diffusion of water molecules in brain tissue, which can be used to infer the orientation and integrity of white matter tracts.

Patients with MMD displayed reduced mean, axial kurtosis and kurtosis, as well as fractional anisotropy in the corona radiata compared to healthy volunteers. Moreover, the posterior limb of the internal capsule showed a decrease in the same parameters in patients with Moyamoya disease. Conversely, axial kurtosis decreased and radial kurtosis increased in the thalami of patients with Moyamoya disease when compared with healthy volunteers (Qiao et al, 2020). DTI has also been used to investigate the relationship between white matter changes and cognitive impairment in patients with moyamoya disease. A study by Liu et al. found that in patients with moyamoya disease the left uncinate fasciculus (UF) and inferior fronto-occipital fasciculus (IFO) could be crucial brain regions that impact arithmetic function, whereas bilateral IFO may influence intelligence (Liu et al., 2020).

Revascularization surgery (RS) is currently the most effective treatment option for both ischemic and hemorrhagic subtypes of the MMD (Acker et al, 2018; Nguyen et al, 2022). The goal of this surgery is to restore blood flow to the brain by creating new blood vessels or bypassing the narrowed or occluded ones. However, at the moment, it still remains underexplored whether cognitive deficit observed in the patients is fully reversible. Recent studies showed that RS was able to effectively alleviate cognitive deficit caused by ischemic stroke in patients with MMD due to restoration of the normal cerebral oxygen metabolism (Yanagihara et al, 2019).

Taken together, researchers have shown growing interest in uncovering the neural underpinnings of impaired cognitive functions in patients with moyamoya disease. However, only a few publications have directly addressed this issue using a combination of fMRI and neuropsychological assessment approaches. In this paper, we present a case study of a patient with moyamoya disease who has been dynamically observed for over four years since the first manifestation of her symptoms.

## 2. Case presentation

A woman in her 30-s was known for her sociable nature and strong-willed character. However, currently, she has lost her ability to perform self-care tasks. In December 2017, she was admitted to the neurological ward of a local hospital due to weakness in her right extremities and speech impairment (see Figure 1 for a graphical representation depicting the full timeline of the case study). A brain MRI revealed that she had an ischemic stroke in the territory of the left middle cerebral artery (MCA). Several months later, in April 2018, she was readmitted to the hospital due to the appearance of left-sided muscle weakness, tetraparesis, and motor aphasia. Neuroimaging results indicated that she had an ischemic stroke in the right parietal and occipital lobes and moyamoya disease (Suzuki stage IV) has been diagnosed. Six months after the initial episode, MRI results revealed the presence of postischemic gliotic and ulegyric changes in the neural tissue of the right temporal and parietal lobes as well as left temporal lobe (Figure 2A). Additionally, small areas of gliotic changes were found in both frontal lobes, as well as multiple focal chronic ischemic changes in the white matter of both hemispheres, especially in the watershed regions. Magnetic resonance angiography (MRA) showed critical stenosis of both internal cerebral arteries in their terminal segments with mild moyamoya vessels at the base of the brain (Figure 2B).

**Figure 1.**
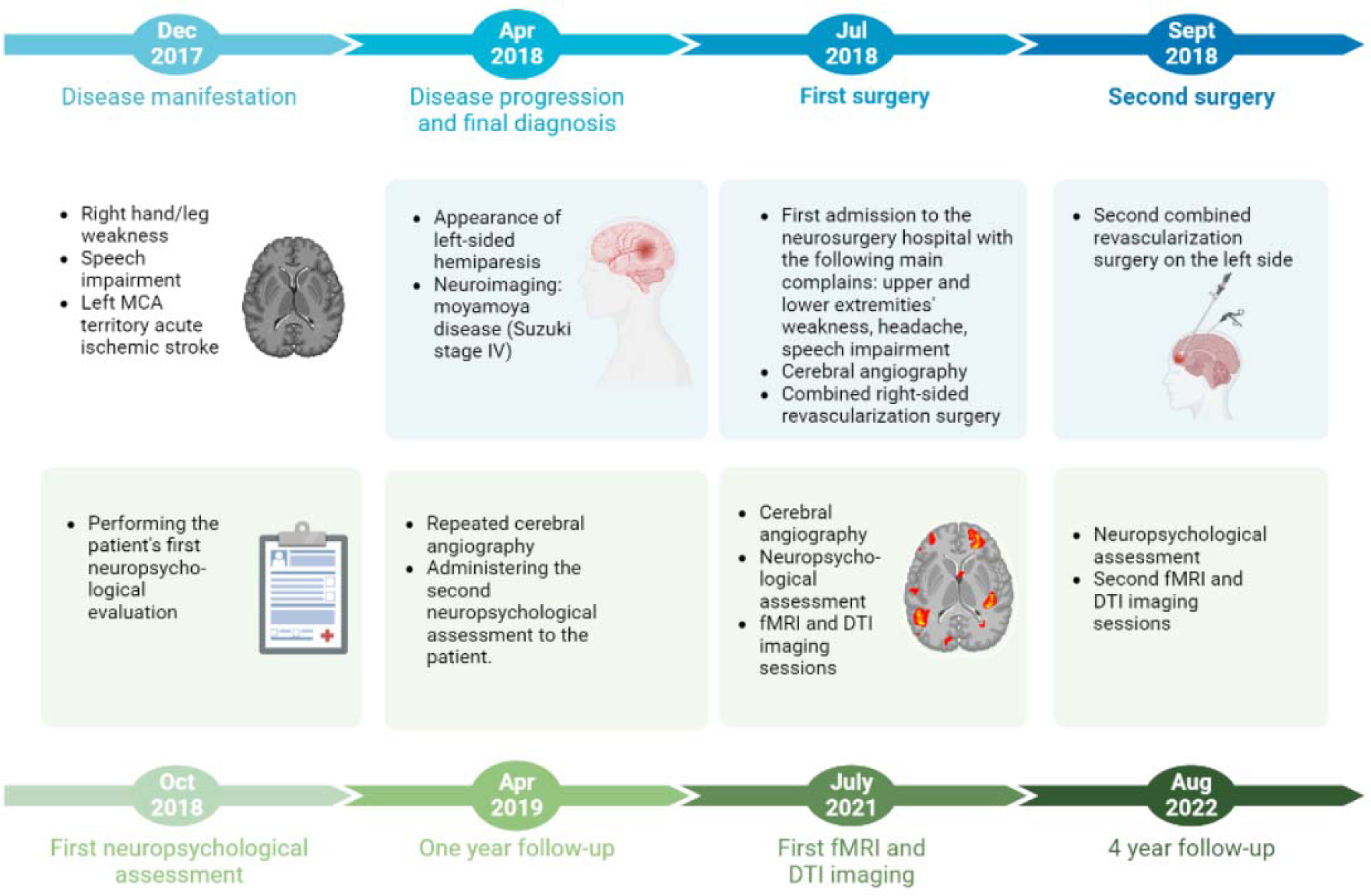
General timeline of the case study.

**Figure 2.**
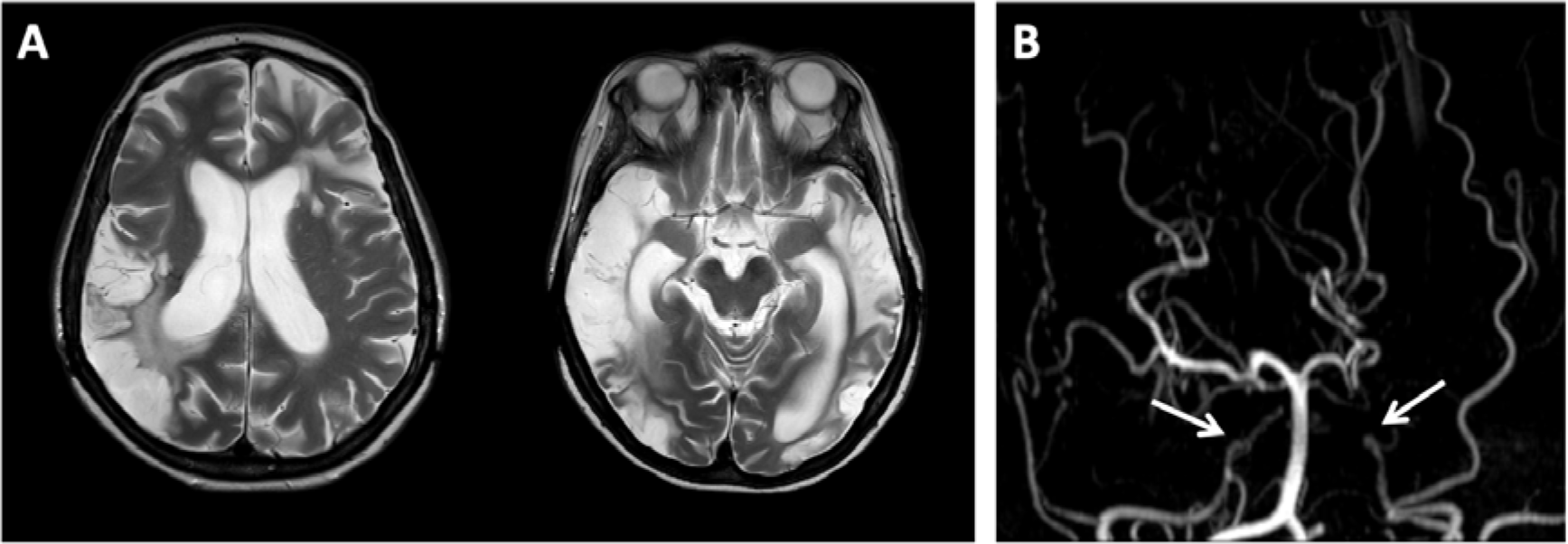
The results of conventional brain MRI (2018). A - Axial T2-WI demonstrates extended bilateral sequelae of ischemic strokes; B - MR-angiography demonstrates critical bilateral stenosis of the terminal segments of ICA (indicated by arrows).

In July 2018, the patient was admitted to our neurosurgical center with complaints of weakness in both upper and lower extremities, intermittent headaches, and speech impairment. The results of digital subtraction angiography (DSA) indicated the occlusion of both internal carotid arteries (ICA) at the level of their clinoid segments (Figure 3) with partial filling of supraclinoids from the posterior circulation. There also was a non-prominent pathological vessel network (moyamoya vessels). These findings were consistent with Suzuki stage 4 of moyamoya disease (Suzuki et al., 1969). The patient underwent right-side combined revascularization surgery which included creation of a double-barreled extra-intracranial microanastomosis between the right superficial temporal artery and M4 segments of the right middle cerebral artery, as well as the creation of encephaloduromyosinangiosis (EDMS) in the frontal, temporal, and parietal areas on the right side.

**Figure 3.**
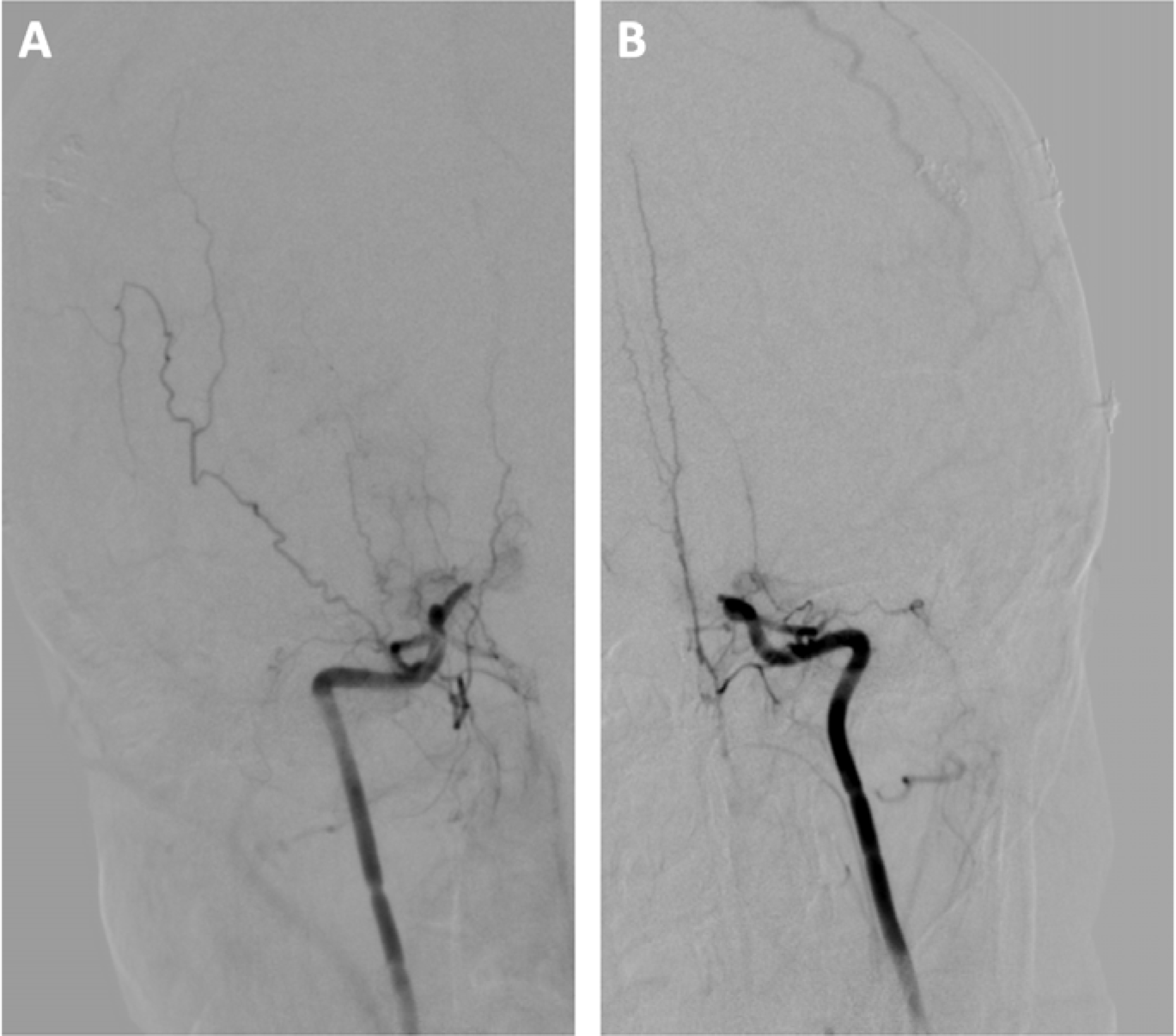
Selective DSA (2018) of right (A) and left (B) internal carotid arteries shows moyamoya disease, stage IV according to the Suzuki classification.

In September 2018, the patient underwent second combined revascularization surgery on the left side. During the surgery, a single-barrel extra-Intracranial bypass was created between the left superficial temporal artery and the M4 segment of the left middle cerebral artery, and EDMS was performed in the frontal, temporal, and parietal regions on the left. Following the second operation, the patient experienced acute postoperative complications and suffered an ischemic stroke that resulted in worsening of speech function deficit.

In April 2019, a control DSA was conducted. The results indicated that the patient had excellent revascularization (grade A according to Matsushima) in the left MCA territory, and good revascularization (grade B) in the right one. There was no increase in stenosis in the posterior circulation system, and the intracranial pathological vascular network was reduced. No new ischemic foci were observed on neuroimaging. In addition, improvement in brain perfusion was noted.

In July 2021, a patient underwent a repeated DSA. Cerebral blood supply reorganization was found with almost complete flow restriction through both ICA, bilateral occlusion of M1 and A1 segments and filling the MCA and ACA circulations from created anastomoses. No new ischemic foci were observed on neuroimaging and cerebral perfusion results were similar to 2019.

### 2.1 Neuropsychological and neurological assessment

Several neuropsychological assessments were conducted using Luria’s qualitative approach to examine cognitive deficits. The assessment included an extensive investigation of memory, attention, perception, and praxis functions. During the first session in October 2018, the patient was unresponsive and unable to follow verbal instructions. The patient did not comprehend spoken language and showed almost no spontaneous speech. The patient’s insight was severely impaired. She demonstrated signs of total aphasia and severe apraxia. Due to these conditions, conducting a full and in-depth neuropsychological assessment was unfeasible.

In April 2019 the patient was tested for the second time. She was available for productive contact, but emotionally labile and exhibited reduced critical thinking abilities. Partial orientation was maintained, as the patient had understood that she was in a hospital in another city and recognized familiar staff members. However, the patient was not oriented in time. Sustained attention was unstable and fluctuated, negatively impacting the completion of diagnostic tests. Productive work periods lasted approximately 30-40 minutes, with attention fluctuations intensifying during periods of fatigue. Partial comprehension of written questions and tasks was possible, with the assistance of visual aids such as signatures on object and plot images. Spontaneous speech was phrasal, with prosodic alterations such as disruptions in pace, rhythm, and intonation. It was also possible for the patient to enumerate established series with visual support.

The patient was able to name objects in images with some verbal and literal paraphasias, as well as copy and independently write words. Phonological, agrammatical, and neurodynamic paragraphias were present, while reading remained intact. Acoustic analysis of rhythmic structures and identification of environmental sounds were inaccessible. There was gross amusia (expressive and impressive) and predominantly sensory aphasia. Visual object recognition was partially preserved (recognizing and correctly naming most presented images), while nonverbal auditory skills were grossly impaired (with a disruption in auditory attention). Partial execution of arithmetic operations within 10 was possible with self-commentary. Kinesthetic (manual, oral-articulatory) praxis was preserved. Objective evaluation of somatosensory recognition was impossible. Body schema assessment showed signs of left-sided neglect.

Constructive spatial activity assessment was impeded due to the severity of frontal syndrome (impairment of planning and voluntary control). This was indicative of regulatory apraxia.

The third neuropsychological assessment was conducted in July 2021. The results showed gross disturbances of auditory gnosis and a mixed systemic speech disorder, primarily characterized by sensory aphasia. During dynamic assessment of the patient’s speech status, she was able to engage in productive contact. However, she was emotionally labile and demonstrated moderately reduced insight. The patient also had partial preservation of orientation and was able to recognize familiar employees of the clinic. Additionally, she was oriented in time and aware that she was in a hospital in another city.

The patient did not react to spoken speech. She was unable to understand oral questions and instructions, but partial understanding of instructions was possible if presented in written form. Her spontaneous speech was agrammatic with gross dysprosody and impoverished oral utterance intelligibility. She was unable to repeat single words or short sentences after the examiner. However, she was able to write, copy words and phrases, with the presence of paragraphias (phonological, agrammatic, neurodynamic). While the patient can read individual words and sentences, understanding the text is difficult. The patient was able to read and partially understood simple written sentences and commands. Her expressive speech was characterized by changes in tempo, rhythm, and intonation, resulting in pronounced dysprosody. She was capable of spontaneously uttering phrases of three to four words. When asked clearly-formulated questions written in block letters, she provided relevant answers. The naming of objects and actions was partially possible. When unfamiliar individuals attempted to draw her attention, she remained indifferent for an extended period. However, she eventually studied their appearance and gestures closely. If the patient became interested in the interlocutor, she attempted to read their lips.

Kinesthetic praxis appeared to be within normal limits without any gross violations. However, the patient experienced difficulties in mastering the motor program, requiring additional presentations and conjugate execution. The learning rate in the experiment was observed to be low. In the trial involving the change of three positions of the brush, the program was simplified for both the right and left hands. The patient showed a violation of coordinated hand movement, specifically reciprocal coordination, and a reduced productivity in constructing the whole from individual elements such as mosaics or Kohs cubes. Additionally, the patient exhibited signs of constructive spatial apraxia, such as confusing the sides of clothes and struggling to button up.

Working with Kohs cubes is unproductive. The individual was able to select the largest and smallest numbers from a set and perform simple counting operations (such as 100-7, 14-7 and 6*2), as well as correctly identify the Roman numeral "16". According to the MMSE, the individual scored 20 points.

During the study of visual perception, it was found that recognition of realistic, crossed-out, and superimposed images, as well as naming colors were not impaired. The patient does not show any signs of neglecting any side of the body or visual field. Additionally, the patient was able to accurately localize touches in her upper and lower extremities.

During the study of the speed of simple and complex sensorimotor reactions, significant impairments were found in attention switching, as well as an inability to perform multiple actions simultaneously. Additionally, difficulties arose when comparing old and new stimuli and providing a response to new signals and their properties, while inhibiting reactions to already familiar stimuli. Correspondingly, rapid exhaustion, decreased scope, and impaired focusing of attention were also observed, as exemplified by the increase in the latency period and the number of errors in serial mathematical actions. Voluntary attention was disrupted, leading to difficulties in switching tasks and affecting the quality of all diagnostic tests. Persistent violations of arbitrary control were present, resulting in numerous errors. Nevertheless, focusing can be enhanced by the presence of interest or increased motivation. As an example, the completion time for the Trail Making Test (TMT) - part A was 1 minute and 35 seconds, while completing TMT-B took over 3 minutes with multiple switching errors.

The patient’s memory functions were also severely impaired. She was asked to reproduce a series of five words. During the first playback, the participant was only able to recall one word, indicating low productivity. During repeated reproductions, the participant’s ability to recall the words showed instability and fragility of imprinting, with a maximum of three words remembered each time in a different combination and order. The participant’s semantic memory was unproductive. When reading a story, the participant experienced difficulties in understanding internal connections and relationships, and was unable to retain and reproduce the general meaning. The participant also failed to answer questions about the text.

The neurological assessment revealed that her fields of vision were not altered, eye movements to the sides, up and down were fully present, but convergence was difficult. The left nasolabial fold was smoothed out, and the tongue deviated slightly to the right. Swallowing was not impaired, phonation was preserved, with moderate dysarthria. Pain sensitivity on the face and limbs was preserved as well. Deep reflexes in the arms were high (higher on the left), and in the legs they were high and equal. Pathological reflexes were present in the left arm. The individual was able to walk independently, with a gait that had elements of hemiparesis. The tone in her left arm and leg was high and it was closer to normal in the right limbs. The left hand was used reluctantly, but movements in the fingers and forearm of the left hand were preserved.

The last neuropsychological assessment was conducted in August 2022. According to her mother, she became more adapted in everyday life (when eating, dressing, performing hygiene procedures, manipulating equipment). She started to react to the sounds of a boiling kettle, a doorbell. For some time after the last hospitalization, she became interested in developmental activities (asked to buy paintings for coloring by numbers, embroidery), paid attention to her appearance, and tried to communicate with friends through social media messaging. Gradually, interest in these activities was lost.

The patient recognized the psychologist she had worked with before, reacted to her with a smile, hugged. She tried to read spoken words by lips, actively responded, and supplemented her answers with gestures. She read the written instructions carefully, commented on them, and confirmed her understanding with a nod. The patient quickly got tired, and there was a gross impairment of attention: TMT-A - 4 minutes 31 seconds, TMT-B - more than 5 minutes with frequent attention-switching errors.

The patient understood written speech (simple sentences and uncomplicated commands). However, she experienced difficulties with complex instructions, such as three-step ones. There were more pronounced grammatical errors and perseverations than before. Expressive speech was altered in terms of speed, rhythm, and intonation (with pronounced dysprosody). In the test of changing hand positions three times, the individual was able to perform the task without errors with both hands, albeit at a slow pace.

Figure 4 summarizes the results of a dynamic analysis of the neuropsychological assessment outcomes for the patient from 2018 to 2022. To present the neuropsychological assessment data succinctly in a single plot, we ranked the degree of functional impairment for each domain on a scale of 0 to 4, where 0 represents intact function and 4 represents complete loss or severe impairment of function.

**Figure 4.**
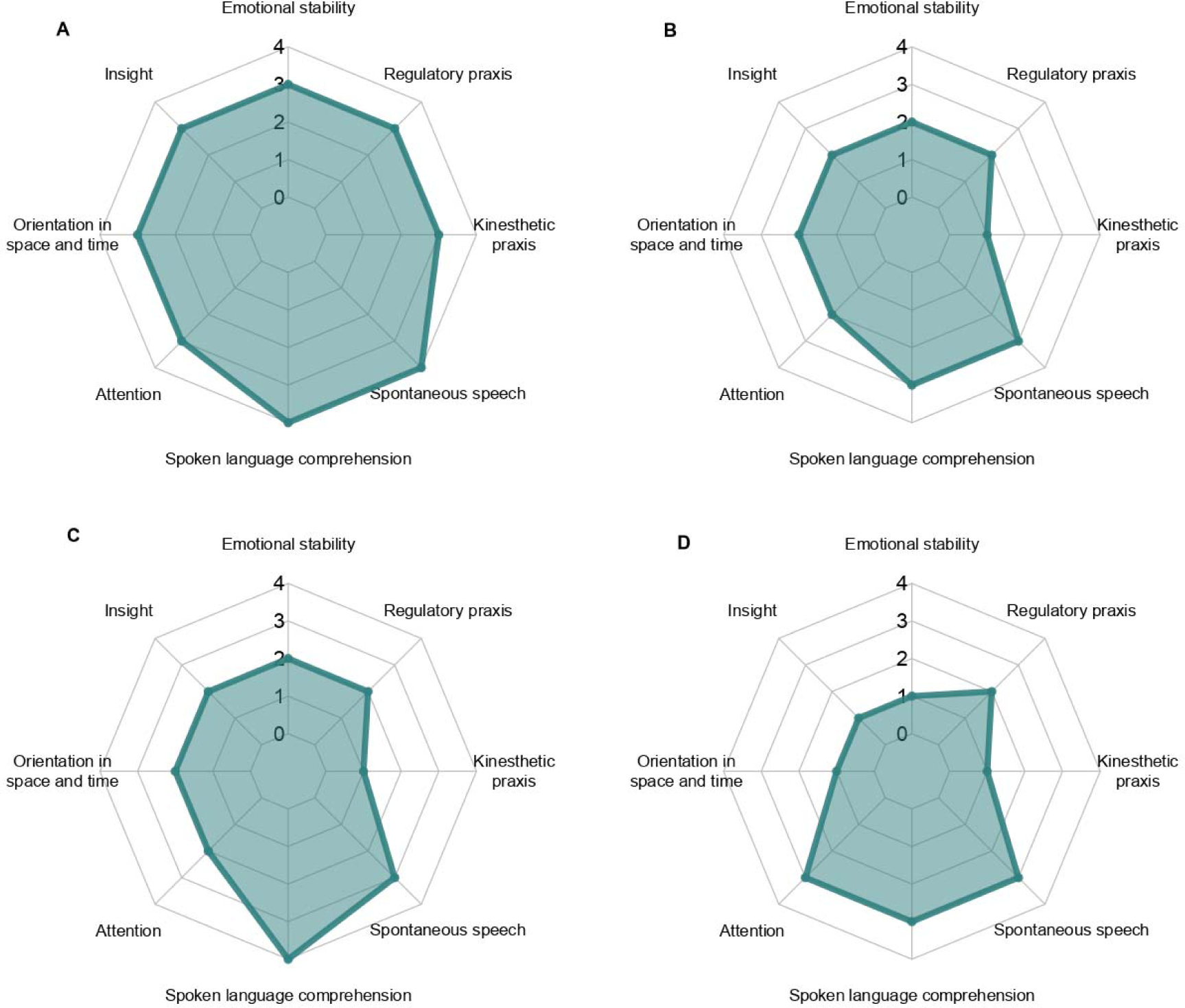
Profile of neuropsychological deficits observed across four sessions: in October 2018 (A), April 2019 (B), July 2021 (С) and August 2022 (D)

### 2.2 fMRI and DTI

Similar to the detailed neuropsychological assessment, functional magnetic resonance imaging and high resolution diffusion tensor imaging were conducted twice, in July 2021 and again in August 2022.

#### 2.2.1. MRI data acquisition and processing

Magnetic resonance imaging was performed using a 3T Philips Ingenia system. The survey included conventional sequences for assessing structural changes (T1-WI, T2-WI, FLAIR, DWI, MR angiography), as well as DTI and functional MRI (both task and resting state).

Task fMRI was conducted in block design paradigm of passive listening to speech (audiobook fragments) and non-speech (music) stimuli, with alternating 30-second periods of rest and listening for 5 minutes, using the following parameters: axial plane, TR – 3000 ms, TE – 30 ms, FOV – 240 mm, Matrix – 80*80, slice thickness – 3 mm, 46 slices with whole brain coverage. Additionally, resting state functional MRI was performed with the following parameters: axial plane, TR – 3000 ms, TE – 30 ms, FOV – 240 mm, Matrix – 80*80, slice thickness – 3 mm, 46 slices with whole brain coverage, 200 volumes. Performing functional studies using more complex paradigms to map language centers was not possible due to the patient’s condition. Diffusion tensor imaging (DTI) was performed with the following parameters: scanning plane - axial, 64 diffusion directions, 3 b0, b=1500; FOV 25.6 cm, matrix 128×128, slice thickness 2.0 mm, 68 slices with whole brain coverage, TR - 8000 msec, TE - 110 msec. The total scan time was approximately 60 minutes.

Task fMRI data was processed using the FSL package (https://fsl.fmrib.ox.ac.uk). The preprocessing pipeline included motion correction, spatial smoothing with a full-width half-maximum of 5 mm, intensity normalization, and brain extraction using FSL BET. No correction of slice time was performed following (Beckmann et al., 2006). To improve anatomical localization of brain activation sites, linear registration of fMRI images on high-resolution T1-VI was performed using FSL FLIRT. Analysis of brain activity in a block design (first-level model-based analysis) was conducted using a general linear model of a set of linear regressions performed separately in the FSL FEAT package, as described by (Monti et al, 2011). Parameters (intensity, area) of the hemodynamic response to activation of certain areas of the cerebral cortex were taken into account.

The resting state fMRI data was processed using the CONN v20b software package (https://web.conn-toolbox.org). The preparation of the fMRI data for analysis included removing the first 10 series of fMRI images, correcting the time of slices, spatial smoothing, correcting geometric distortions, co-registering on structural images, and registering in the standard coordinate system (MNI) with a resolution of 3 mm^3^. The connectivity between regions of interest (ROI) and all other brain regions was analyzed voxel-wise. The ROI was selected based on existing literature data, where the nodes of the default mode network (DMN) and speech network were used for connectivity analysis (Table 1).

**Table 1.** The MNI coordinates of the region of interest were analyzed during the assessment of the individual’s connectivity profile.

| Brain network | ROI | MNI coordinates |
| --- | --- | --- |
| Default Mode Network | Posterior cingulate cortex (PCC) | 1; -61; 38 |
|  | Medial prefrontal cortex (MPFC) | 1; 55; -3 |
|  | Inferior parietal lobule (LP, R&L) | 47; -67; 29 (R)<br>-37; -67; 29 (L) |
| Speech Network | Inferior frontal gyrus (L) | -51; 21; 15 |
|  | Superior temporal gyrus (L) | -54; -51; 30 |

DTI data processing was conducted using FSL (https://fsl.fmrib.ox.ac.uk/fsl) and Explore DTI software (https://www.exploredti.com/) and included motion correction, correction of geometric distortions, co-registration with structural images, calculation of fractional anisotropy maps. Tracts reconstruction performed manually with TrackVis (https://trackvis.org/), and a probabilistic algorithm was utilized. Such tracts as arcuate fasciculus, uncinate fasciculus, inferior fronto-occipital fasciculus, inferior longitudinal fasciculus, and frontal aslant tract were reconstructed

#### 2.2.2 Results

##### 2021

###### Task fMRI

After task fMRI data processing and analysis, a single cluster of activation was discovered in the posterior third of the superior temporal gyrus in the left hemisphere in response to speech stimuli (audiobook fragments). This region is recognized as the site of the auditory-motor interface, which was previously referred to as the Wernicke speech center.The cluster was found within the unchanged brain tissue and was reduced in size. No other activation clusters were detected (Figure 5).

**Figure 5.**
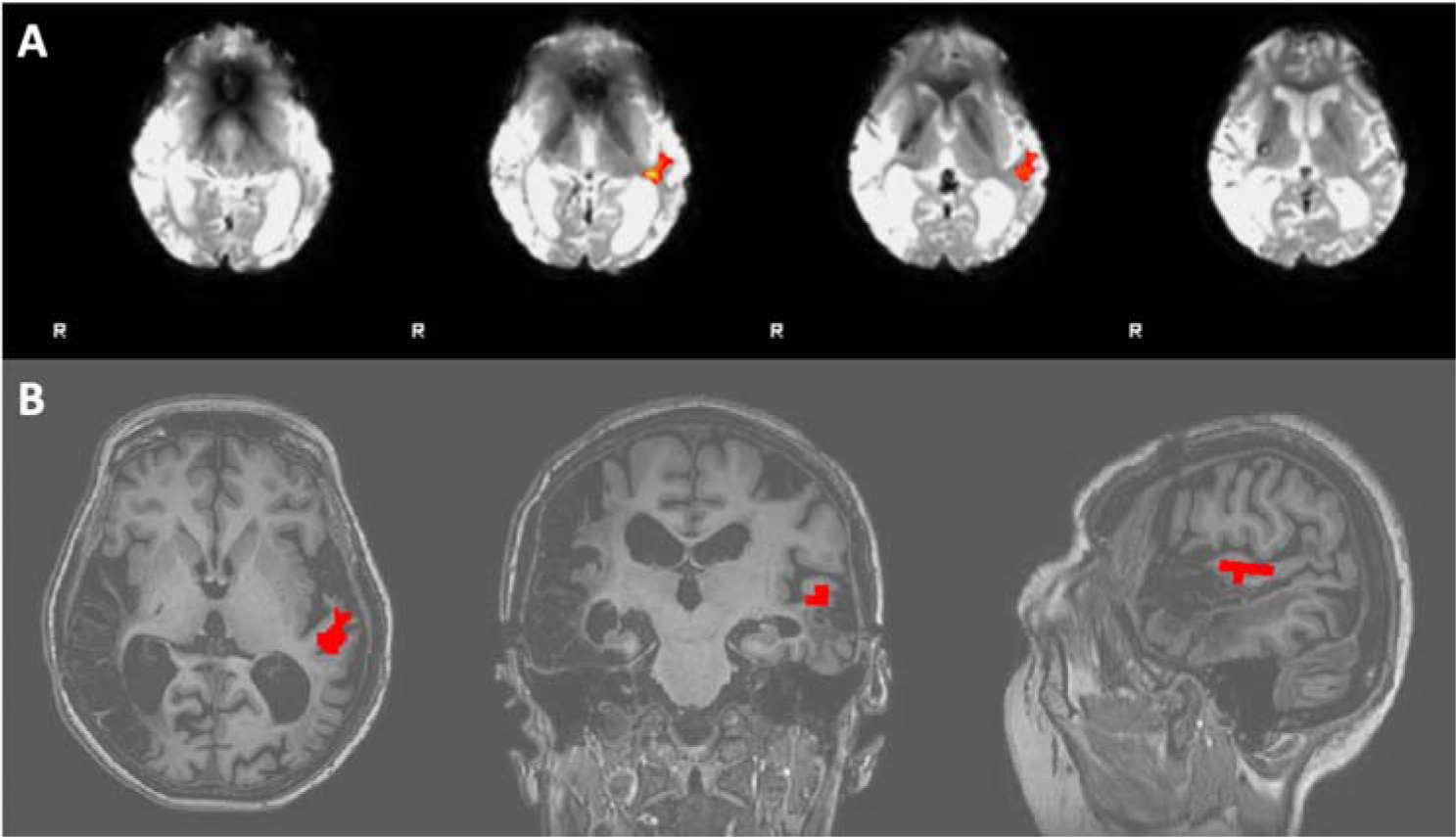
The results of task fMRI during passive listening of speech stimuli. A - FSL FEAT analysis results; B - activation cluster in posterior part of left superior temporal gyrus after coregistration with T1-WI.

During the fMRI paradigm of passive listening to non-speech stimuli, multiple activation clusters were identified (Figure 6). These clusters included the left pre- and post-central gyri, as well as the right hemisphere of the cerebellum. Furthermore, activation was also detected in the middle third of the right cingulate gyrus. Additionally, there was activation observed in the poles of both temporal lobes.

**Figure 6.**
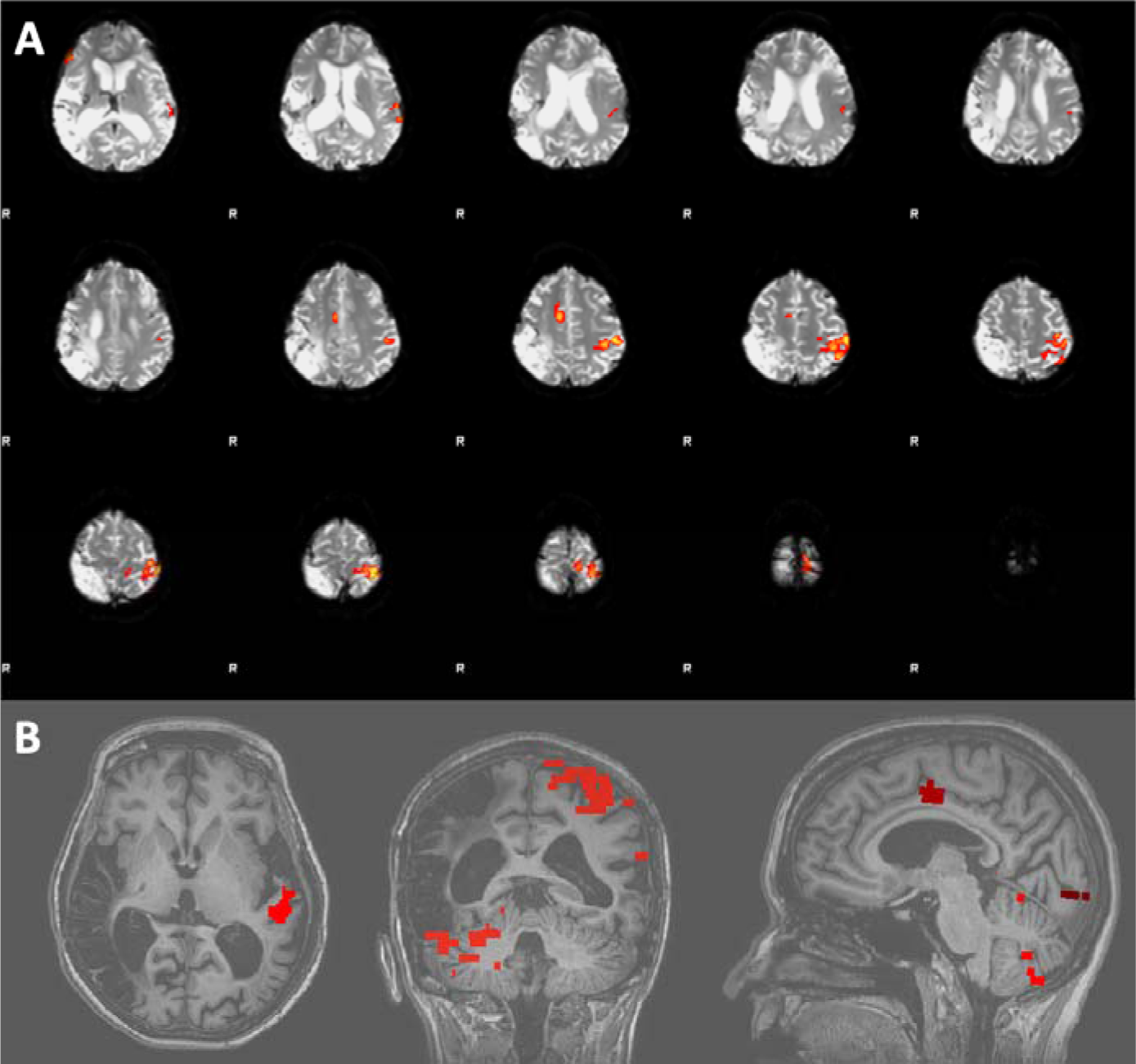
The results of task fMRI during passive listening of music stimuli. A - FSL FEAT analysis results; B - multiple activation clusters after coregistration with T1-WI.

###### Resting state fMRI

The patient’s resting state fMRI profile was analyzed using seed-based analysis. The main nodes of the Default Mode Network (DMN) and their connectivity with other functional regions of the cortex were visualized (Figure 7). Our findings indicate that there was preserved connectivity between the mPFC, PCC, and right parietal lobule. However, postischemic changes were noted in the left inferior parietal lobule, resulting in almost complete loss of functional activity in this area.

**Figure 7.**
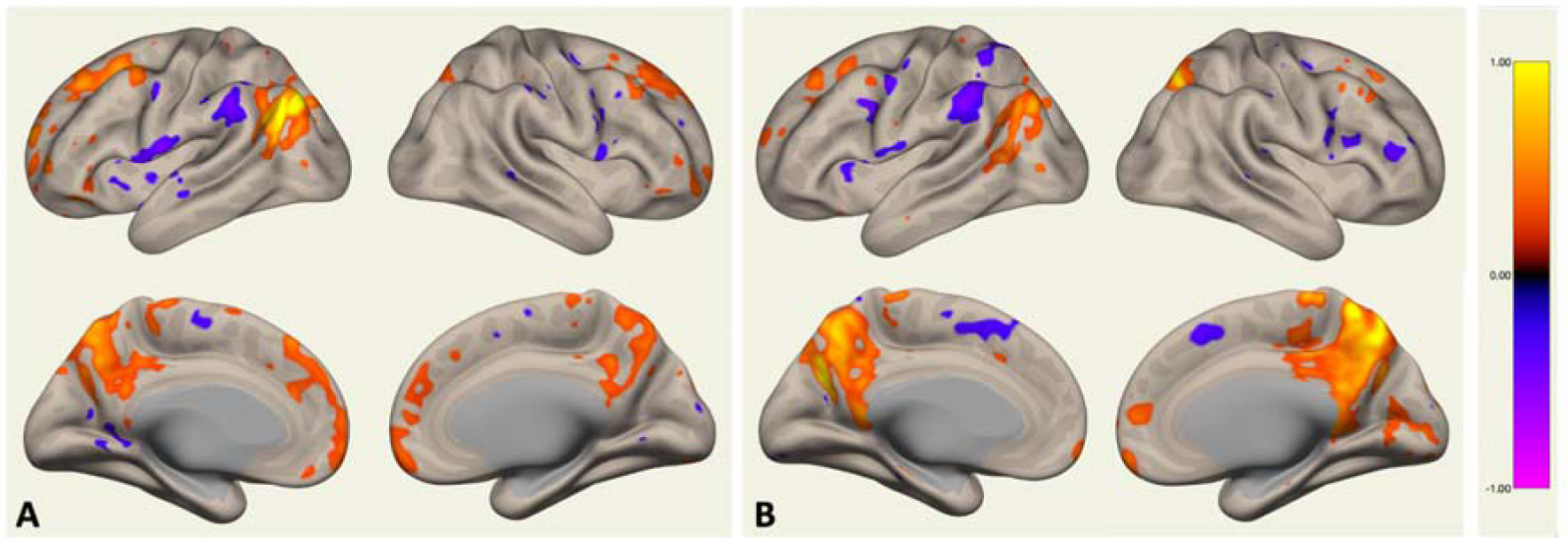
The results of resting-state fMRI seed-based analysis (Default Mode Network): A - the seed is in the mPFC, B - the seed is in the PCC. The primary nodes of DMN were preserved, except for the left parietal lobule.

Additionally, the main nodes of Language Network (posterior part of the left superior temporal gyrus - pSTG, and inferior frontal gyrus - IFG) were visualized when seeds were chosen in the left hemisphere (Figure 8). However, we detected the disconnection between dorsal and ventral streams of the language network (the nodes of dorsal stream were not activated when seed was in the pSTG and the nodes of ventral stream were not activated when seed was in the IFG). Furthemore, the nodes of lexical phonological network and ventral stream’s semantic nodes (which are normally localized in the middle and inferior temporal gyri of both hemispheres), as well as dorsal stream’s articulation nodes (in the lateral parts of precentral gyri and supplementary motor cortex bilaterally) were not visualized.

**Figure 8.**
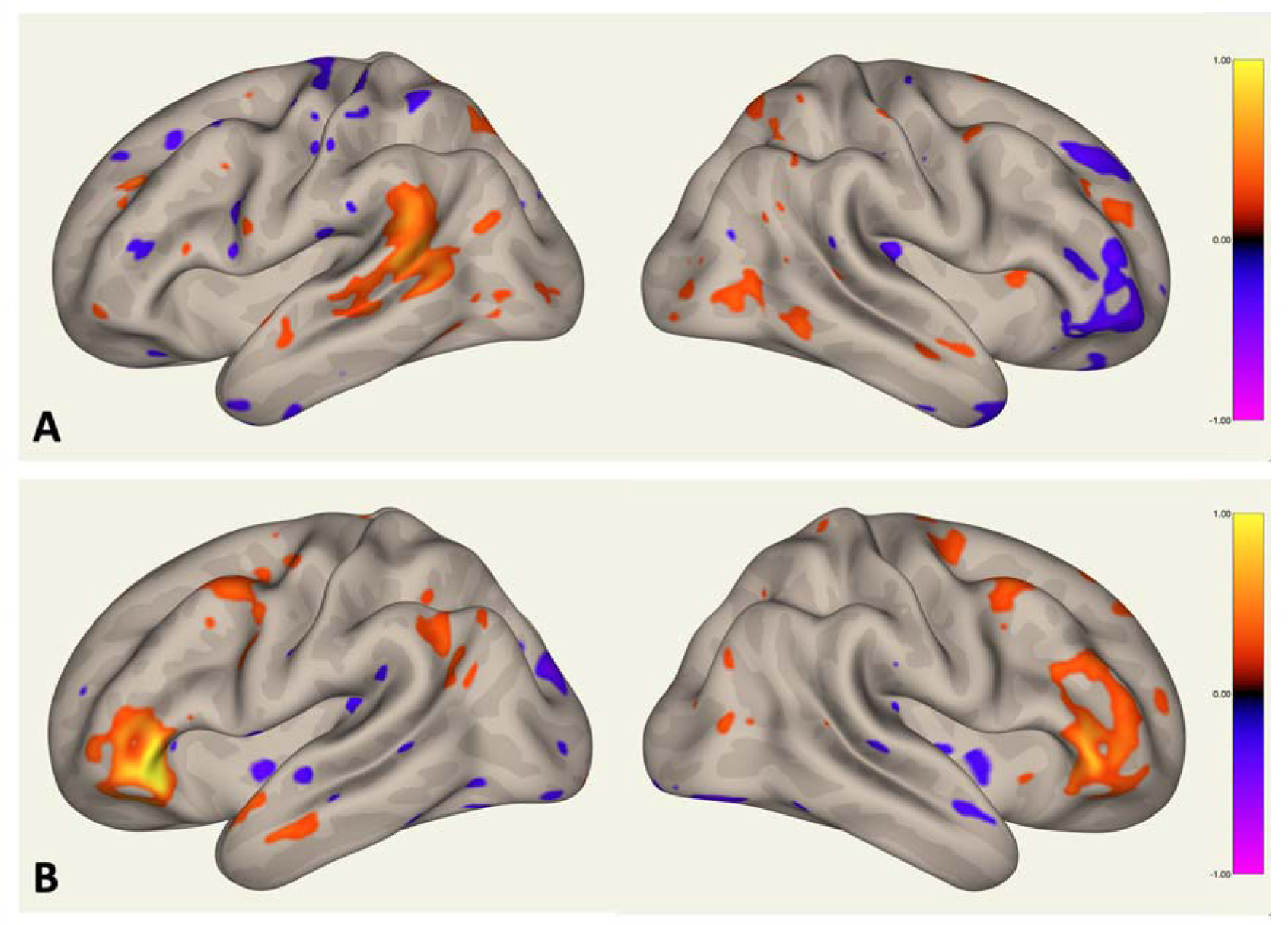
The results of resting-state fMRI seed-based analysis (Language Network): A - seed is the posterior part of the left superior temporal gyrus; B - seed is the left inferior frontal gyrus. There is disconnection between dorsal and ventral streams of the network (the nodes of dorsal stream are not activated when seed is in the ventral stream and vice versa); additionally, the nodes of lexical phonological network and ventral stream’s semantic nodes, as well as dorsal stream’s articulation nodes are not visualized.

###### DTI (with MR-tractography)

MR-tractography was used to reconstruct the main speech-related tracts of the dominant hemisphere (Figure 9), including the arcuate, inferior frontal-occipital, inferior longitudinal, uncinate, and aslant tracts. While the anatomy appeared to be preserved, the tracts themselves were thinned and the coefficient of fractional anisotropy was decreased (FA = 0.38-0.4).

**Figure 9.**
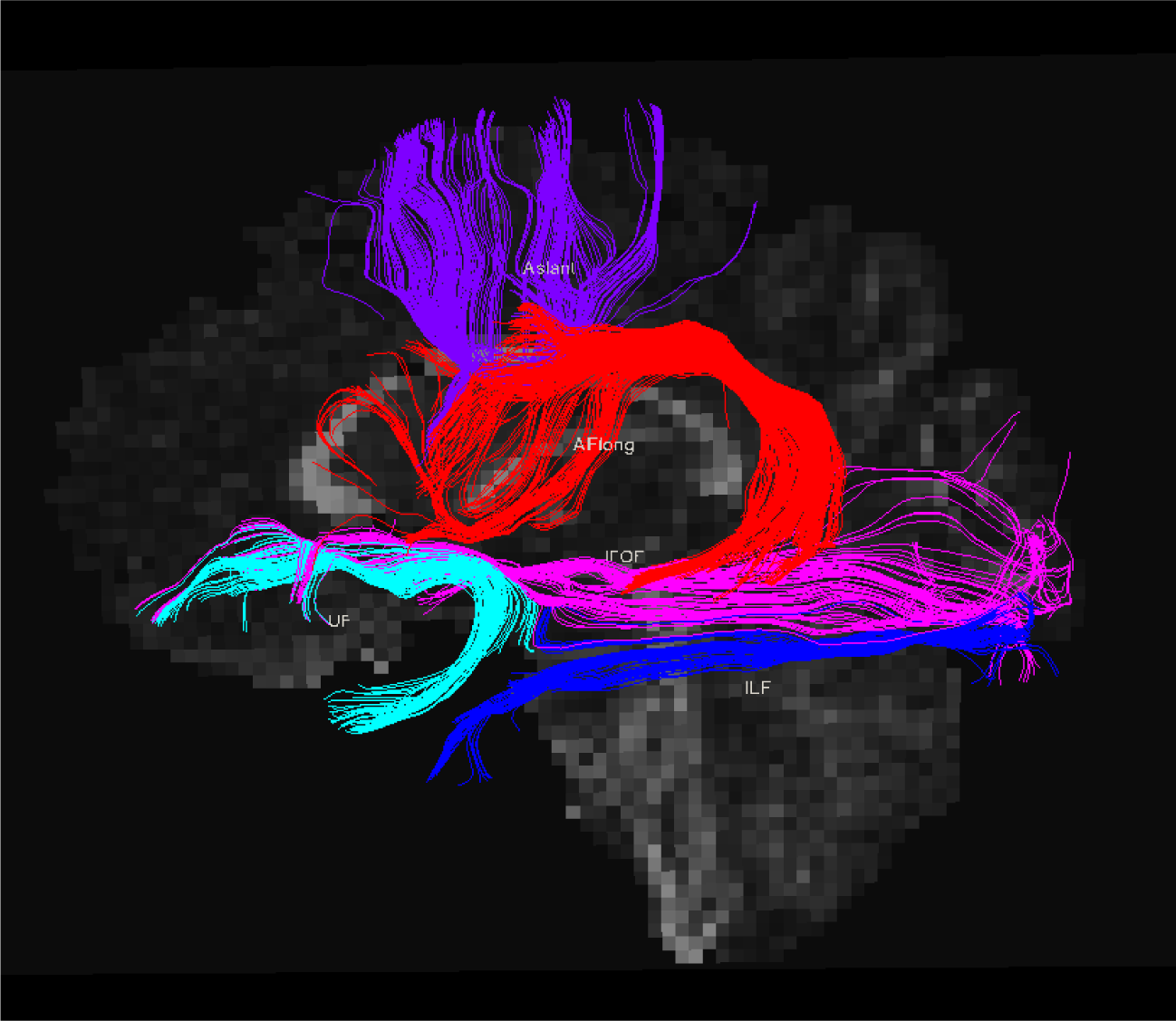
Patient’s arcuate fasciculus (red), aslant (violet), uncinate (turquoise), inferior longitudinal fasciculus (bright blue) and inferior fronto-occipital fasciculus (pink)

##### 2022

In general, the results of both tractography and functional MRI analyses were consistent with those obtained in 2021. While the anatomy of the tracts was preserved, there was evidence of thinning and impairment of structural integrity. During passive listening to audiobook fragments, we observed a single activation cluster located at the auditory-motor interface in the posterior third of the left superior temporal gyrus. This cluster had decreased in size with an activation threshold set at Z=5 and no other clusters were identified during this portion of our study. However, when participants listened passively to music stimuli, multiple activation clusters emerged including those within the left pre- and postcentral gyrus, right cerebellum, middle third of right cingulate gyrus as well as bilateral temporal poles.

The individual connectivity profile (according to resting state fMRI data) was also similar to session in 2021 (not shown).

## 3. Discussion

Despite being a rare disease, the study of moyamoya disease holds significant importance as it can provide valuable insights into the mechanisms of cerebrovascular diseases and assist in developing new treatments for related conditions. This article is, to the best of our knowledge, the first instance where longitudinal tracking of cognitive functioning and its neuronal underpinnings was conducted on a single patient with moyamoya disease over a period of four years.

We found that significant speech impairments in our patient were reflected in altered structural and functional brain architecture. The extensive areas of gliotic changes in brain tissue in both temporal lobes as a result of multiple strokes were revealed according to conventional MRI data. It is well known that the temporal cortex contains the number of eloquent regions that participate in language processing, such as nodes of the lexical phonological network and the semantic nodes of the ventral stream (Hickok et al., 2007). In this study, only one neuronal activation cluster was detected in the posterior third of the superior temporal gyrus in our patient with task fMRI upon listening to speech stimuli. Moreover, DTI analysis revealed a notable thinning of the main language-associated white matter tracts in the left hemisphere. Furthermore, the disconnection between the dorsal and ventral streams of the language network was detected, as well as the lack of activation of the nodes of the various language networks according to the resting state fMRI. Interestingly, despite multiple gliotic changes, the general DMN architecture and connectivity pattern were preserved.

Speech impairments associated with moyamoya disease have been studied much less extensively than other cognitive changes. During the fMRI study of auditory perception in this patient, activation clusters were detected in the left pre- and postcentral gyrus, right hemisphere of the cerebellum, right midcingulate cortex, and bilateral temporal poles when listening to non-speech stimuli (music). The obtained results are consistent with the literature data on the involvement of the aforementioned areas in processing both speech and non-speech stimuli. Cerebellum is now known to be a part of a distributed network devoted to auditory stimuli processing generally and speech stimuli in particular (Petacchi et al., 2005; Knolle et al., 2012). Interestingly, in our study, we did not observe any increases in cerebellar activity during the perception of speech stimuli. This finding contrasts somewhat with available data in this research field where studies consistently report associations between speech processing and cerebellar activation (Silveri, 2021). Right midcingulate cortex activation may reflect emotional processing of the stimuli as this region is believed to be involved in emotion regulation broadly (Pereira et al., 2010; Caruana et al., 2018). Finally, when listening to musical stimuli, activation areas are often detected in the temporal poles. Different authors interpret these findings either as evidence of the involvement of these regions in processing emotionally charged information or as the implementation of semantic processing of auditory stimuli (Koelsch, 2005; Belfi and Tranel, 2014).

During speech stimuli perception (listening to audiobook fragments), only a reduction in the size of the activation zone was observed within Wernicke’s area for our patient. Additionally, resting-state fMRI did not reveal any functional connections between regions belonging to dorsal and ventral network streams which could significantly contribute to language impairments identified in this patient. We also found a significant thinning of the main white matter tracts involved in language processing in our patient. Reduced fractional anisotropy values in arcuate fasciculus add to the evidence of changed Language network properties in this patient, which is in line with the results of an accumulated body of studies investigating structural characteristics of white matter tracts in patients with speech impairments (Marchina et al., 2011). Earlier studies in patients with moyamoya have repeatedly demonstrated the presence of white matter changes heavily impacting their cognitive functioning (Kazumata et al., 2015). For instance, different diffusivity metrics have been shown to be altered in anterior thalamic radiation, inferior fronto-occipital fasciculus, superior longitudinal fasciculus, uncinate fasciculus, inferior longitudinal fasciculus in these patients (Calviere et al., 2012; Liu et al., 2020). At the same time, we were unable to find any mention in the scientific literature regarding changes in the arcuate fasciculus among patients with moyamoya disease. This may potentially be due to the fact that speech impairments are often mild or absent in these patients, and other cognitive deficits tend to dominate their clinical presentation.

Beyond that, prominent disturbances in a range of cognitive and behavioral domains like orientation, insight, reasoning abilities, praxis, attention, body schema were revealed as well. The results presented here are partially consistent with published literature, which suggests that patients with moyamoya disease exhibit persistent dysfunction in the organization of spatiotemporal patterns of neural activity (Kazumata et al., 2017; Liu et al., 2021). Previous studies have demonstrated significant reorganization of intra- and internetwork connectivity profiles in patients compared to healthy controls (Lei et al., 2016; Lei et al., 2020). For example, DMN functional connectivity was altered in these patients but restored to resemble that of healthy controls after undergoing vascular reconstruction surgery (Sakamoto et al., 2018). However, our study did not identify disrupted connectivity within the DMN. The main seeds of DMN-mPFC, PCC, and right parietal lobule - retained their functional connections. Nonetheless, we observed postischemic changes in the left inferior parietal lobule resulting in almost complete loss of functional activity in this area. This might have contributed to a relative disconnection between the anterior and posterior nodes of the default mode network. One possible explanation for not detecting gross violations in the functional architecture of DMN may be attributed to the fact that previous studies were conducted on samples consisting of dozens of patients while ours is a single-case study.

Cognitive impairments in moyamoya disease occupy a significant place in the overall structure of observed symptoms, seriously reducing the quality of life for such patients. In full accordance with previously published data, our patient demonstrated marked memory impairments, decreased sensory-motor reaction speed, and regulatory praxis disturbances while the kinesthetic praxis was relatively preserved (Festa et al., 2010). During dynamic observation over 4 years, notable improvements were found in the patient’s insight, ability to orient herself in time and space, personal identity as well as social functioning. However, when investigating attention we found discrepancies with Festa et al.’s findings who did not identify significant attention deficits in moyamoya patients whereas our patient displayed marked attention deficits throughout the observations (Festa et al., 2009). It should be noted that other research groups have indeed reported similar deficits in patients (Fang et al., 2016; Shi et al., 2020).

Revascularization surgery remains an effective therapeutic method for moyamoya disease, providing prevention of potential strokes (Ando et al., 2019). The most effective approach involves the combined use of direct and indirect methods, known as combined revascularization. The direct component provides immediate effects on blood flow to the brain, protecting against stroke immediately after surgery, while the indirect components promote further long-term cerebral blood supply. Recent studies have convincingly demonstrated that revascularization surgery has the ability to restore blood flow and white matter parameters, reorganize profiles of pathological functional activity in patients’ brains leading to restoration of cognitive function (Kazumata et al., 2019; Gao et al., 2022; Yasuda et al., 2022). Following two surgeries performed in 2018, our patient has not shown any signs of recurrent strokes for at least 4 years. Additionally, there has been partial restoration of her cognitive abilities and social skills. Specifically, we observed improvements in our patient’s speech comprehension abilities, ability to spontaneously generate words and short sentences, moderate improvements in their written text comprehension and ability to count within ten. However, even at the four-year follow-up, our patient continued to exhibit persistent and marked impairments in all of the above-mentioned functions.

It should be added that this study has several limitations worth mentioning. Firstly, the neuropsychological assessment was conducted in accordance with Luria’s approach, which places a strong emphasis on qualitative data processing rather than quantitative data. This approach may hinder direct comparisons of both intra-(over multiple time points) and inter-subject performance levels in the different cognitive domains tested. Furthermore, the neuropsychological assessments at different follow-up points were conducted by different specialists, and there was no unified protocol available. This can also be considered a limiting factor in the final interpretation of the results. Additionally, we did not have the patient’s fMRI and DTI data for the period immediately preceding the manifestation of her first symptoms. Finally, although we cannot completely rule out the possibility of spontaneous patient’s cognitive status improvement, which may not be directly related to surgery performed, revascularization surgery remains the gold standard for the restoration of impaired brain’s blood flow in patients with moyamoya disease.

Taken together, despite significant improvements after the revascularization surgery, our patient still demonstrates marked cognitive deficits that directly impact her quality of life. However, the results of this study indicate the necessity and utility of dynamic observation of patients to reflect on changes in symptom severity and improvement trajectories resulting from therapy.

## Informed consent

Patient signed an informed consent form for publishing this data.

## Credit author statement

P.A. - manuscript draft preparation and editing, F.E. - neuroimaging and manuscript editing, M.A. - neuropsychological assessment, O.K - surgery, G.G. - language skills assessment, M.G. and R.J. - general design, supervision, manuscript editing.

## Data Availability

All data produced in the present study are available upon reasonable request to the authors

## Abbreviations

MMD: - moyamoya disease
ICA: - internal carotid artery
MRI: - magnetic- resonance imaging
DTI: - diffusion tensor imaging
RSN: - resting state network
CBF: - cerebral blood flow
RS: - revascularization surgery
EDMS: - encephaloduromyosinangiosis

